# Diabetes-related Distress and the Association to Hypertension and Cardiovascular Disease Among Individuals Living with Type 2 Diabetes in Rural areas in Vietnam

**DOI:** 10.1101/2023.02.06.23285554

**Authors:** Amalie Sophie Sahl, Diep Khong Thi, Thanh Nguen Duc, Dieu Huyen, Jens Søndergaard, Janni Nielsen, Ib Christian Bygbjerg, Tine Gammeltoft, Dan W. Meyrowitsch

## Abstract

**Objective:** The prevalence of diabetes has been rising in rural areas of Vietnam over the last years to the extend where it has become a public health burden. Individuals with diabetes-related distress (DRD) is in greater risk of adverse health outcomes e.g. lower blood sugar control and polypharmacy. The objective of this study is to assess the association between hypertension and cardiovascular disease (CVD) and the occurrence of DRD among individuals with type 2 diabetes (T2D) in rural areas of Vietnam.

**Method:** This is a cross-sectional study of 806 individuals who had been receiving treatment for T2D at a district hospital in the northern Vietnamese province Thai Binh. Based on self-reported data DRD was assessed through Problem Areas in Diabetes scale 5 (PAID5) and defined as a score of 8 or above. The occurrence of the comorbid conditions hypertension and CVD were self-reported.

**Results:** Among 806 individuals with T2D 37.7% of the men and 62.3% of the women presented with DRD. Out of the total group 35.6% reported hypertension, 7.3% reported CVD and 21.2% reported a combination of hypertension and CVD. The results of the multivariate analyses showed that the odds ratio of DRD was significantly higher (OR=1.67, CI95: 1.11-2.52) in the group who reported a combination of hypertension and CVD.

**Conclusion:** Among individuals with T2D in rural areas of Vietnam there is an increased risk of DRD if a combination of hypertension and cardiovascular disease is also present. Hence, considering diabetes-related comorbidities can be useful in order to successfully identify individuals in risk of DRD.

## BACKGROUND

Over the last decades the occurrence of type 2 diabetes (T2D) has increased rapidly throughout Asia with three-fold to five-fold increase in several countries suggesting that by 2025 as many as 333 million people will live with T2D[1]. In 2012, the prevalence of diabetes in Vietnam was 5.4% with a lower prevalence of diabetes in the rural communities as compared to urban communities but is now on the rise and the increasing number of individuals living with T2D is becoming a financial and public health burden[2, 3].

Diabetes-related distress (DRD) is defined as the emotional distress and the behavioural changes associated with diabetes[4, 5]. This includes feeling overwhelmed, scared and angry when thinking about diabetes, as well as not feeling motivated to maintain diabetes self-management[6].

DRD is common among individuals with T2D. A systematic review, including 55 studies from different countries, showed that approximately 36% of individuals with T2D were suffering from DRD[7]. In a previous study on DRD, with data from the same population as used in the present study, 50% of the individuals reported symptoms of DRD[8]. In addition, a smaller study from a medical centre in Vietnam showed that approximately 12% of the individuals diagnosed with T2D presented with DRD[9].

Besides the psychological impact of DRD, studies from high-income countries report that DRD influences glycaemic control and self-care: people with DRD had higher blood levels of triglycerides and higher BMI[5, 10].

Additionally, two more recent studies based on analyses of data from the same population in Vietnam as used in the present study, DRD was associated with the occurrence of polypharmacy and unmet needs for social support[8, 11].

Given the adverse health outcomes associated with DRD, it is important to consider and address the risks of developing DRD in the planning and implementing of new interventions directed towards the treatment of T2D. This will increase both the quality of care and the relevance of this care for individuals living with T2D[5].

Comorbidities are common in individuals living with diabetes. Individuals with diabetes have an increased risk of cardiovascular disease (CVD) and this risk increases further if hypertension is also present[12]. A systematic review showed that CVD globally affects 32.2% of individuals living with T2D[13], and in the US occurrence of ischaemic heart disease is higher among individuals with T2D who showed symptoms of DRD [14, 15].

An improved understanding of risk factors for DRD, and in turn the possibility of identifying individuals at risk of DRD, and therefore those who may have a higher risk of diabetes-related complications can potentially inform the development of effective and cost-effective treatment. This is specifically aimed at individuals diagnosed with T2D who are living in resource-constrained communities where treatment is challenging.

Therefore, a cross-sectional study was conducted to assess the association between hypertension and/or CVD and DRD among individuals living with T2D in Thai Binh Province, Vietnam.

## METHODS

### Study Design, Setting, Recruitment and Sample

A cross sectional survey was carried out in Thai Binh Province Vietnam from December 2018 to Febuary 2019. Thai Binh Province is a coastal province located in the northern part of Vietnam. The province is divided into 7 districts and one city and covers an area of 1542 km^2^ and consisted of a population of approximately 1.8 million individuals in 2019. Self-reported data were collected as part of the research project “Living Together with Chronic Disease: Informal Support for Diabetes Management in Vietnam (VALID)”.

The sample included individuals who had been diagnosed with T2D after the age of 40. The age of diagnose criteria was set in order to exclude individuals who could possibly have type 1 diabetes.

Based on district hospital records, two districts in Thai Binh Province were randomly selected for the study. The two districts were Quynh Phu District and Vu Thu District in the northern part and the southern part of the province, respectively. Quynh Phu District and Vu Thu District include two district hospitals and one district hospital. In each district the researchers selected the two communes with the highest number of individuals with T2D, as well as two neighboring communes where data collection was easily accessible, resulting in four selected communes from each district. Hence, a total of eight communes were purposely selected for the present study.

A total of 963 individuals who were treated for T2D in the district hospitals in the eight communes were initially invited to be included in the cross-sectional survey. Among these, 37 (3.8%) refused to participate and 78 (8.1%) did not stay at the address reported to the hospital or had moved away at the time of data collection resulting in a total of 848 people who participated in the survey. Additionally, 42 individuals were excluded from the final sample, as they either had been diagnosed with diabetes prior to the age of 40 or did not remember when they were diagnosed. The final study sample included a total of 806 individuals diagnosed with T2D.

### Data Collection and Quality Control

From each of the eight selected communes two village health workers were engaged in data collection. The village health works were responsible for the data collection in their own commune. Prior to the survey the interviewers attended a course consisting of a two-day workshop including testing and subsequent revision of the questionnaires. This was followed by one day of field-based training.

The interviewees were invited to participate in the survey either by mobile phone calls or by personal visits. Each participant was informed about the purpose of the interview in writing and verbally and, if agreeing in participation, asked to fill in a form of consent. The interviews were conducted individually and face-to-face in the participants homes to secure proper understanding of the questions as well as discretion in questions containing sensitive matters.

The survey questionnaire included a total of 176 questions with eight areas: the participants and their households, health and use of healthcare services, the participants lifestyle, homework and management of diabetes, informal social support, sexual wellbeing and emotions, use of smartphones and digital media, informal support, emotional impact, need for support, and use of electronic devices and internet. In the present study, demographic and socio-economic information about the participants, their households, their lifestyles and their health, was used. To ensure correct translation to Vietnamese and back to English the questions were pre-tested and subsequently revised prior to data collection among a group of Vietnamese people with T2D. This was done in order to ensure the optimal understanding and interpretation of the questions and respective answering options.

### Outcome Variable

DRD was assessed using the 5-item Problem Areas of Diabetes Distress (PAID5), a short form of PAID that has earlier proven good validity as a short form measure of DRD[16]. The PAID5 assess measures of DRD through five questions regarding emotional distress in relation to diabetes. In each question the patients were asked to what degree they had experienced the relevant feeling within the last four weeks prior to the interview. Items were scored on a 4-point Likert Scale, from 0 (not a problem) to 4 (serious problem) resulting in a possible total score from 0 to 20. A total score of ≥8 points was defined as an indication of DRD[16]. To ensure validity and correct translation and understanding the Vietnamese PAID5 questionnaire was tested in a group of Vietnamese individuals with diabetes. Prior to the pilot-testing PAID5 had translated into Vietnamese and translated back to English.

### Exposure Variables

As part of the questionnaire the participants were asked whether they had been diagnosed with hypertension, epilepsy, depression, liver and kidney disease, bone and joint problem, CVD or any other comorbid disease. For this analysis three physical comorbid predictors were chosen: 1) hypertension; 2) CVD; and 3) a combination of both hypertension and CVD.

### Co-variables

Demographic and socio-economic characteristics were chosen as covariables based on previous literature, including a recent study of this present sample[8], and a priori based on an assumption that these variables could function as predictors for DRD in this specific socio-cultural context (Appendix E). Gender was categorized into: 1) Male; and 2) Female. Age was categorized into four age groups: 1) 40-49 years; 2) 50-59 years; 3) 60-69 years; and 4) ≥70 years. Current occupation was categorized into: 1) Unemployed; 2) Farmer; 3) Employment in small trade, as worker, as government employee, in a private company; and 4) Retired. The economic situation was based on self-reported household income and divided into: 1) Poor; 2) Near poor; 3) Medium; and 4) Wealthy. Relationship status was divided into: 1) Single; 2) Married; 3) Divorced; and 4) Widowed. Household size was categorised according to number of individuals in the household: 1) Living alone; 2) 2 people; 3) 3 people; and 4) ≥4 people. Social support of relevance for T2D was divided into: 1) No unmet need for emotional support; 2) Unmet need for emotional support. Levels of physical exercise were based on how often the individuals exercised for 30 minutes or more: 1) Never or rarely; 2) Weekly to monthly; 3) Daily. Educational status was categorized according to highest achieved education: 1) No school; 2) Primary school; 3) Highschool; 4) University or above. Smoking status was categorized into: 1) Never smoked; 2) Previously smoked and; 3) Present smoker.

### Statistical Analysis

A descriptive analysis was performed, and results were presented as frequencies for the total sample, for the group of individuals presenting with DRD and as prevalence of DRD in each group. Co-variables which had a statistically significant level of association with the outcome variable in the bivariate analyses using a significance level of p<0.05 were included in the subsequent multivariate analyses. Results were presented as odd ratios (ORs) with 95% confidence interval (CI 95%). Further, the association between co-variables with statistically significant p-values and DRD are shown as ORs with CI 95%. To assess risk of effect modification, stratified analyses for all co-variables were performed with Breslow-day tests for homogeneity of ORs across strata, with a significance level of p<0.05 for all co-variables. SPSS Statistic Software Package (version 28) was used for data analysis.

## RESULTS

### General and Socio-economic Characteristics of Individuals with Type 2 Diabetes

Characteristics of the total sample and distributions of individuals with DRD are presented in Table 1. As we have been reporting in a previous study[8] we found that among the 806 individuals with an age of 40 or above, 50.0% (n=403) presented with DRD. Approximately half of the included individuals were female (52.7%) and found that being female had a statistically significant association with DRD (OR: 2.17, CI95%: 1.64-2.88). The majority were married and lived with one or more family members. Most individuals had an educational level of primary school or above, a moderate income and were engaged in daily physical activities. Majority of the individuals (88,3%) in the sample were non-smokers. With regards to education level and occupational status, the highest prevalence of DRD was observed among unemployed individuals (64.3%) and individuals who had no school education (70.0%). The prevalence of DRD among individuals who identified as being either poor or near poor were 78.7% and 67.6%, respectively. The percentage of individuals who reported an unmet need for emotional support of relevance for their diabetes self-management was 7.7%. In this group of individuals, the prevalence of DRD was 77.5% as compared to 48.6% among individuals who did not report an unmet need for emotional support. There was a statistically significant difference in prevalence of DRD across groups with various levels of physical activity with the lowest prevalence observed among individuals who exercised (44.7%). Furthermore, there was a statically significant difference in prevalence of DRD across groups with different smoking status with the lowest prevalence among present smokers (31.9%).

**Table 1.**
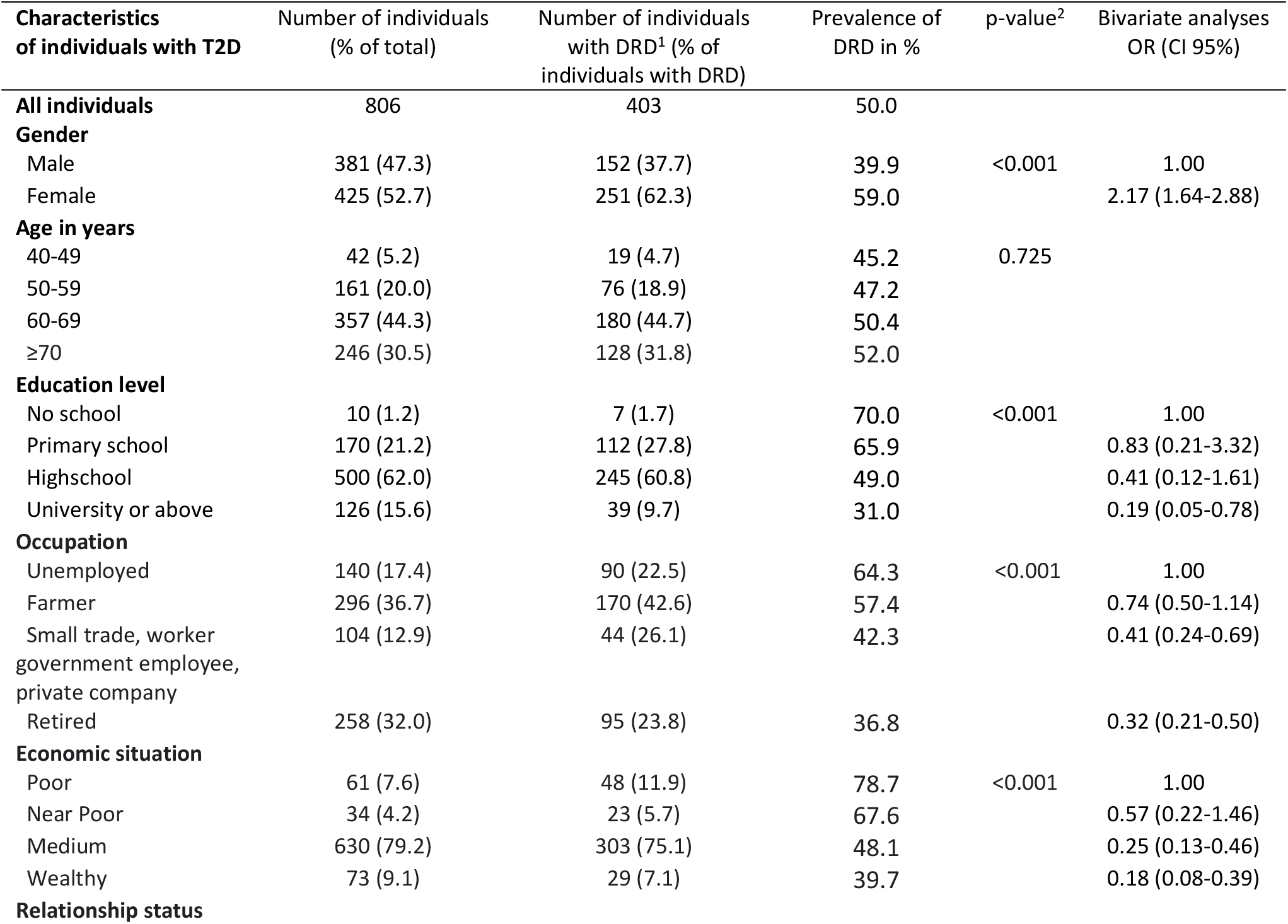
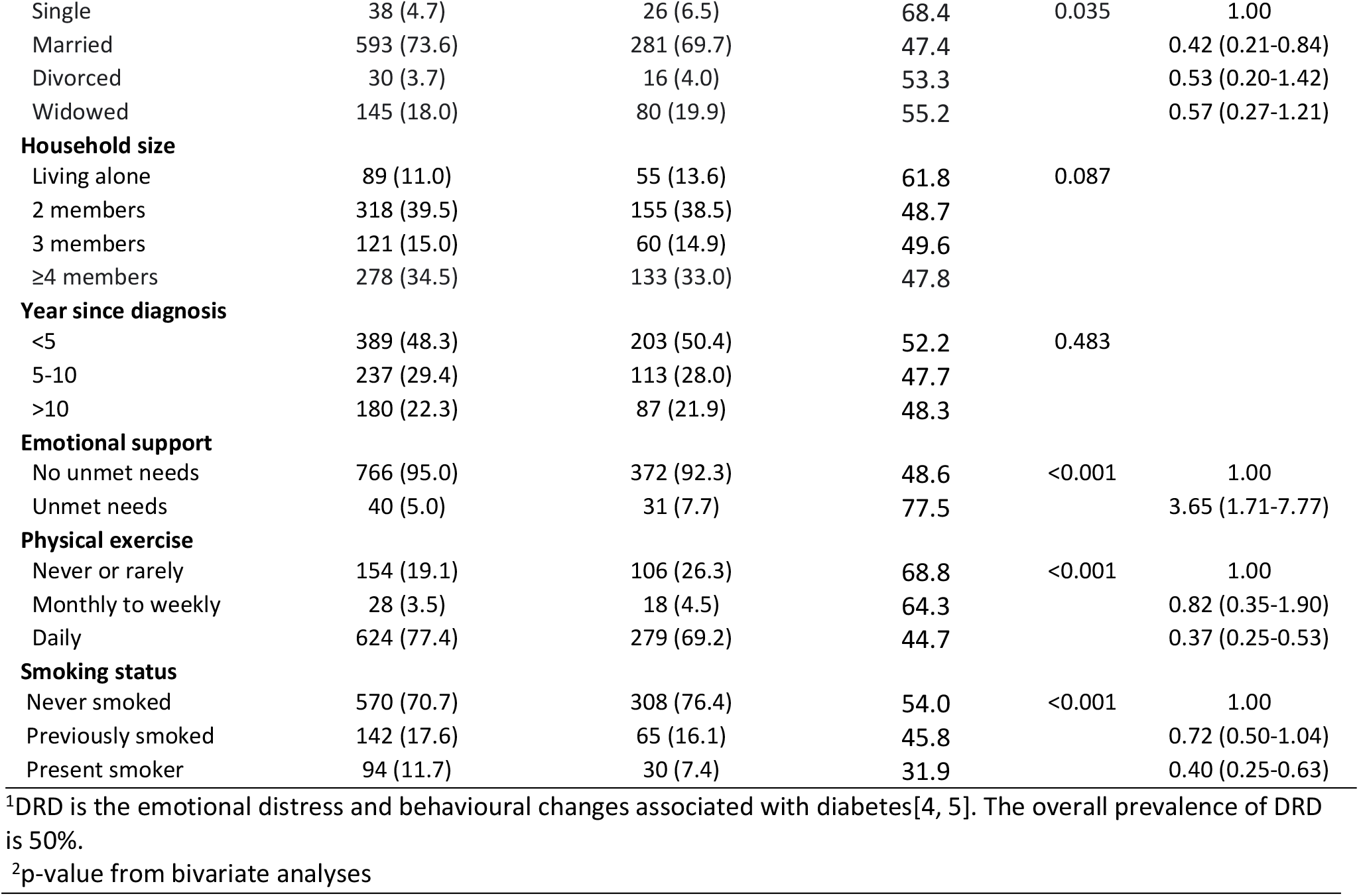
Demographic and social characteristics of the studied group with type-2 Diabetes.

We found that gender, education level, occupation, economic situation, relationship status, emotional support, level of physical exercise and smoking status were statistically significant predictors of DRD.

### Impacts of Self-reported Hypertension and Cardiovascular Disease on Diabetes Distress

Among individuals with T2D, 35.6% and 7.3% reported only hypertension and CVD as comorbidities, respectively, and additionally 21.2% reported a combination of hypertension and CVD (Table 2). Results of the analyses of the associations between comorbidities and DRD among individuals living with T2D are presented in Table 2. The results of multivariate analyses showed that individuals with a combination of hypertension and CVD had a higher odds ratio for DRD as compared to individuals with T2D only (OR=1.67; CI95%: 1.11-2.52). There was no significantly increased odds ratio for DRD among individuals with hypertension alone or CVD alone and DRD (OR=1.17; CI95%: 0.81-1.64 and OR=0.92; CI95%: 0.50-1.70, respectively).

**Table 2.** Results of bivariate and multivariate analyses of the associations between the selected predictors, hypertension and CVD, and DRD.

| Characteristics Of individuals with disease | Number of individuals (% of total sample) | Number of individuals with DRD (% of total sample) | Prevalence of DRD in % | Results of statistical analyses |  |
| --- | --- | --- | --- | --- | --- |
|  |  |  |  | Bivariate analyses<br>Crude OR (CI95%) | Multivariate analysis<br>OR (CI 95%)<sup>3</sup> |
| TD2 only | 289 (35.9) | 135 (16.7) | 46.7 | 1.00 | 1.00 |
| TD2 + Hypertension<sup>1</sup> | 287 (35.6) | 143 (17.7) | 49.8 | 1.13 (0.82-1.57) | 1.17(0.81-1.64) |
| TD2 + CVD<sup>2</sup> | 59 (7.3) | 27 (6.7) | 45.8 | 0.96 (0.55-1.69) | 0.92 (0.50-1.70) |
| TD2 + Hypertension + CVD | 171 (21.2) | 98 (12.2) | 57.3 | 1.53 (1.05-2.24) | 1.67 (1.11-2.52) |

<sup>1</sup>Individuals with CVD removed from analyses
<sup>2</sup>Individuals with hypertension removed from analyses
<sup>3</sup>Adjusted for gender, education, occupation, economic situation, relationship status, emotional support, physical exercise and smoking status

### Possible Effect Modification of Co-variables

The results of Breslow-day tests for analysis of possible effect modification are shown in Table 3. The co-variable age functioned as a statistically significant effect modifier in the association between hypertension only and DRD (χ2=10.16, df=3, p=0.0017); and physical exercise functioned as a statistically significant effect modifier in the association between the group that presented with a combination of hypertension and CVD and DRD (χ2=7.137, df=2, p=0.028). No effect modification was observed for other co-variables (Stratified Odds Ratios is available in appendix A, B and C).

**Table 3.**
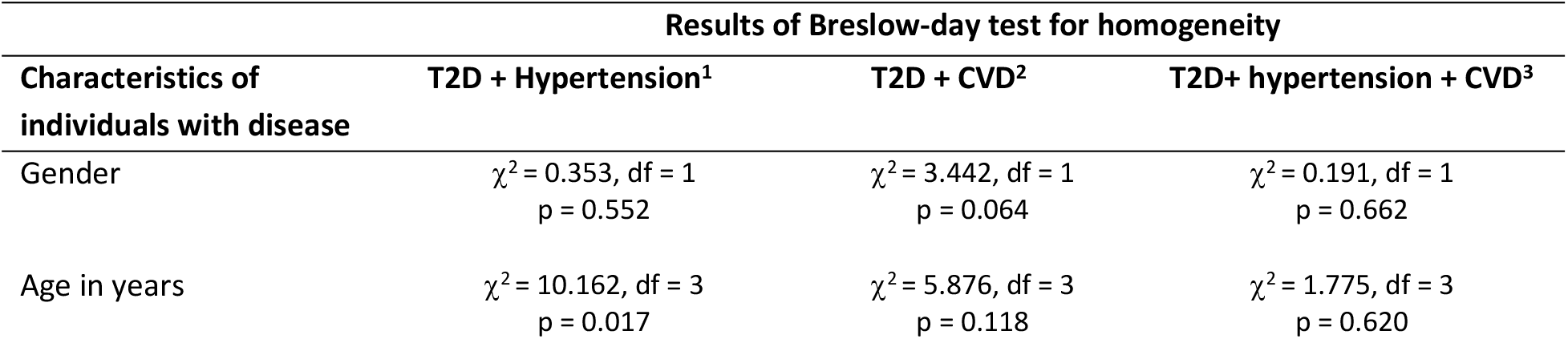
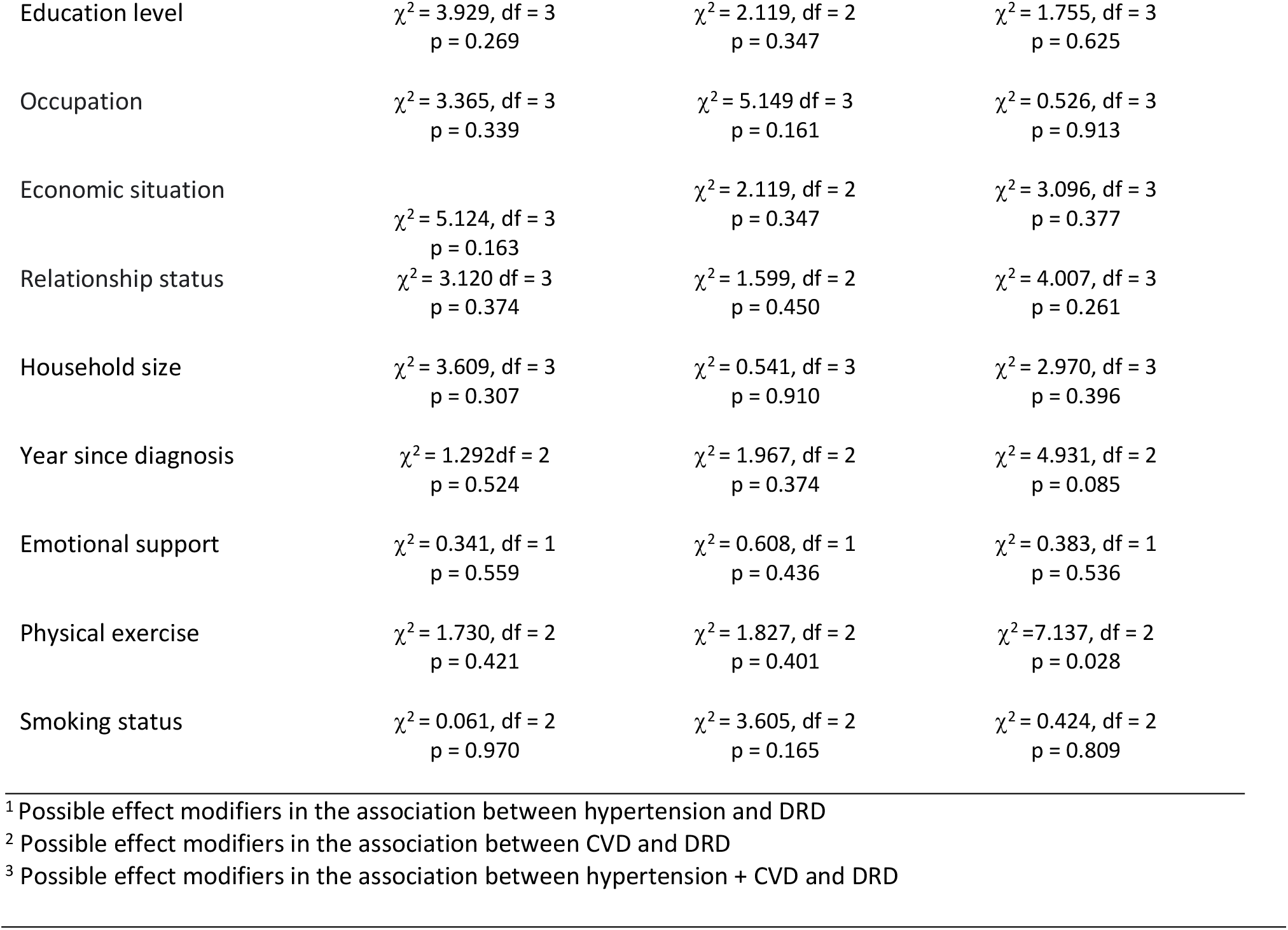
Breslow-day tests for homogeneity of the odds ratios within the comorbidity subgroups of the studied group with type-2 diabetes

## DISCUSSION

In the present study, the prevalence of DRD were 39.9% among the men and 59.0% among the women, respectively (Table 1), and a combination of hypertension and CVD was a statistically significant predictor for DRD (Table 2).

In comparison with the observed prevalence of DRD among individuals living with T2D in the present study, a lower prevalence of DRD was reported from a study in Ho Chi Minh City, Vietnam (29.4%)[17]. However, large differences in prevalence of DRD across Asian countries have been reported with 8.9 % in Thailand, 42.2% in China, 49.2% in Malaysia and 76.2% in Pakistan [18-21]. These differences might be caused by variations in the populations studied regarding demographic and socio-economic factors, access to and quality of healthcare, study designs and tools for assessment of DRD. To assess DRD we used PAID5 while Diabetes Distress Scale (DDS) was used in other studies [17-21]. Though both PAID5 and DDS are considered valid tools for assessment of DRD[22], PAID5 specifically covers the emotional aspects of DRD while DDS focuses on diabetes self-management, food-related problems and stress related to treatment[22]. This might contribute to the difference in DRD prevalence between the studies. PAID5 has not been validated in Vietnam but it has been validated in internationally as a reliable tool for quick assessment of DRD with a sensitivity of 94% and a specificity 89% of for detecting DRD [16]. Furthermore PAID5 is has been validated as a reliable assessment tool in other Asian countries including China[23] and Korea[24].

In the present study, being female was associated with an increased risk of DRD. This finding corresponds to the finding of a large meta-analysis including studies from the USA, the Netherlands China and Singapore[7]. In Vietnam, after marriage women are more likely to settle down in the household of the husband’s family, resulting in separation from their own family[25], which potentially could isolate the women from their natal relatives. This could lead to lack of emotional support and potentially lead to a higher risk of DRD amongst females later in life. Another aspect of the gender-specific practices in Vietnam is that women are more often responsible for taking care of the household and all the members, both children and the elderly[25]. This practical and emotional responsibility could additionally contribute to a higher level of DRD.

Several studies have found that being a present smoker was associated with DRD[26, 27]. In the present study, we found the lowest prevalence of DRD among smokers, followed by earlier smokers and the highest prevalence of DRD among the individuals who never smoked. Since DRD in our study was self-reported, the difference in results compared to previous studies could be due to underreporting of DRD within the group of smokers, which might have caused a differential misclassification. Additionally, in Vietnam it is uncommon for female individuals to smoke. A Vietnamese study reported that the prevalence of current smoking among male and female patients in health care facilities were 34.6% and 1.1%, respectively[28]. This difference in distribution between gender may likely contribute to our results.

T2D and hypertension are often present at the same time due to multiple physiological factors[29]. This corresponds well with the results of our study where we found that over half of individuals (56.8%) reported having hypertension with an additional 21.2% reporting having a combination of hypertension and CVD. The high proportion of individuals who reported CVD in addition to hypertension could be caused by the fact that many are diagnosed late or have poor blood pressure control as reported in a previous Vietnamese study [30] leading to cardio-vascular complications. Only 7.3% reported having CVD as only comorbidity. This may be due to the fact that CVD is overrepresented among individuals with both T2D and hypertension[12].

A previous study, which included the same sample as used in the present study, reported an association between number of unmet needs for social support, including emotional and economic needs, and the risk of DRD[8]. This suggest that individuals presenting with DRD are not only at risk of a more severe course of disease but are also in a social perspective a more exposed group, which makes them important to identify in order to successfully aim the treatment towards T2D in rural Vietnam. The previous study further reported that the risk of developing DRD increased with number of types of unmet needs for social support[8]. The increased risk of DRD among individuals with multiple unmet needs aligns with the results in the present study. In the present study, the risk of DRD increased if the individual reported multiple comorbidities, suggesting that the risk of DRD is associated with an increased amount of pressure. The result of the present study showed that presenting with a combination of hypertension and CVD in combination with diabetes statistically increased the risk of DRD. This is supporting the assumption that T2D comorbidities are important to take into consideration in order to predict variation in individual risk of a more severe course of disease and a poorer treatment compliance. This assumption is further supported by a study from Thailand[18] and a study from Iran [31] that both showed an association between diabetic comorbidity and the risk of DRD. Since the cross-sectional nature of the design does not allow us to determine the direction of causalities, the association between DRD and individuals with multiple comorbidities could also be caused by the physical complications which commonly co-exist with DRD, e.g. poorer blood sugar control and higher levels of triglycerides[5, 10] causing a higher prevalence of individuals with comorbidities.

Presenting with only hypertension as a diabetic comorbidity showed a positive association to DRD, though it was not a significant predictor. Several earlier studies have linked hypertension to mental unease, including a study carried out in rural areas of Vietnam that showed that quality of life among individuals living with hypertension was low regarding the psychological aspects of health[32], and a German study that found that in individuals with T2D hypertension reduced quality of life[33]. This adds to the assumption that there could be an association between hypertension and mental well-being, including DRD, within individuals with T2D, but this will need further investigation in order to assess in detail.

There was no increased risk of DRD in the CVD subgroup. This contrasts with earlier studies showing that CVD increased risk of DRD among individuals living with diabetes mellitus [14, 15, 34]. The observations in the present study could, however, be due to the very small sample of individuals only reporting CVD. Hence, lack of statistical power would reduce the chance of detecting a statistically significant association between CVD and DRD.

The present study included a relatively large sample size and had a high participation and response rate. The study had, however, some limitations. Though our study included a large sample size, the comorbidity subgroups had pronounced differences in sample size.

In the present study, participants were recruited through lists of individuals diagnosed and treated at the district hospitals. Individuals treated at province hospitals or national referral hospitals were not included in the sample. In Vietnam, individuals with more severe disease or in need of more treatment are often referred for treatment at the province or national referral hospitals. This will most likely lead to a selection bias and exclude individuals with more severe T2D related complications and comorbidities. Also, this study was carried out in Thai Binh Province and the findings may not be representative for other areas of Vietnam.

Self-reported questionnaire-based data is a great representative of self-perception. Nonetheless, a part of the Vietnamese practise includes a strong wish of not troubling or being a burden to others and, hence, a strong tendency to tone down personal problems and obstacles.(Gammeltoft, Tine M: The Force of Love: Type II Diabetes in Vietnam as Tentatively Transformative Experince), [35]. This could potentially lead to an underreporting of DRD and hereby a reporting bias which could potentially lead to differential misclassification.

Furthermore, since data on comorbidities was based on self-reported information, these exposure variables may also be prone to various types of misclassifications. This could lead to an underestimation of reported comorbidities, as the likelihood of identifying with a disease increases with the severity of the disease and the need for treatment, thereby potentially leaving out the milder cases. In contrast, a Chinese study showed that having DRD has a negative impact on self-reported health status[36], indicating that there might be an overreporting of comorbidities. If the misclassification of comorbidity data were evenly distributed between individuals with and without comorbidity this non-differential misclassification would, however, not affect the observed strength of the associations between hypertension and/or CVD and DRD.

## CONCLUSION

The findings in the present study suggest that in rural areas in Vietnam there is an increased risk of DRD among individuals who reported a combination of hypertension and CVD, compared to those who live with T2D and none of the comorbidities. The findings of this study suggest that taking diabetes-related comorbidities into consideration when treating T2D can potentially be of great importance in order to successfully identify individuals at risk of DRD, and thereby improve diabetes treatment and outcome. Further studies with more participants are needed in order to fully understand the relationship between comorbidities and DRD.

## Data Availability

Additional data used for analyses in this current study are available from the corresponding author upon request.

## Acknowledgement

We are grateful to the research assistants from Quynh Phu District and Vu Thu District who conducted the interviews, and we thank the staff at the Population Center and Commune Health Centers in Thai Binh Province for their assistance. Additionally, we would like to thank everyone from Quynh Phu District and Vu Thu District who participated in the interviews. We also want to thank the Biostatistics department of University of Copenhagen for providing assistance with the Breslow-day analyses.

## Author Responsibilities

All authors contributed to subsequent revisions and editing of the manuscript.

